# Bacterial Contamination of Surgical Site Wounds Among Post-Operative Patients and Theatre Surfaces at Hoima Regional Referral Hospital

**DOI:** 10.64898/2026.07.09.26357612

**Authors:** Abarteneko Chelangat, Winnie Akiteng, John Roberts Padde, Musanad Asimai, Moses Tabule, Jimmy Omeke, Rudolf Buga, Bwaga Ibrahim

## Abstract

Bacterial contamination of Surgical sites could lead to Surgical Site infections (SSI) which may prolong hospital stay, increased treatment costs and increased mortality. This study determined the prevalence of bacterial contamination of surgical sites among post-operative wounds and theatre surfaces together with their resistance to commonly prescribed antibiotics. A cross-sectional study design was used where a total of 290 and 74 swabs were collected from patients and theatre surfaces respectively. Swabs were cultured on duplicate plates of Blood Agar, Chocolate Agar and MacConkey Agar. Gram-staining and Biochemical tests were performed to identify the isolates. Resistance to commonly prescribed antibiotics was determined using the Kirby Bauer (KB) method. Data were analyzed using SPSS version 23, and descriptive statistics, Chi square and student T-tests were used to describe the results. The prevalence of bacterial contamination in wounds was 30.7% and was significantly higher in women of child bearing age (χ2= 10.79, df=1, *P=0*.*0010*). Microbial growth increased with an increase in duration of antibiotic therapy (χ2=12.73, df=2, *P=0*.*007*). *E. coli* was responsible for the highest cases of wound contamination (34.9%). All microorganisms isolated from post-operative wounds showed considerable resistance to antimicrobials. All isolates from wounds were resitant to Trimethoprin Sulfamexathone and 76.9% showed resistance to Ciprofloxacin. Other than *E*.*coli* and *Acinetobacter*, the rest of the isolates were susceptible to imipinem. Fourty nine gram positive isolates were grown from theatre surfaces and a significant majority (86%) were from air. There was high resistance to Erythromycin in Coagulase Negative *Staphylococcus* (CNS) isolates (56.0%). Overall, our study demonstrated that wound contamination at the Hoima Regional Referral Hospital is high but not associated with theatre surface contamination.

## 1. Introduction

Wound contamination is one of the major causes of prolonged hospital stays and blood sepsis. Wounds are majorly contaminated with bacterial organisms that colonize the skin and those that inhabit hospital surfaces, and the quantity and type of these organisms varry by the type of wounds(Ashoobi et al., 2023; Deku et al., 2024). Wounds types may be presented as acute surgical wounds, post surgical wounds, chronic woulds including diabetic wounds and traumatic wounds like those from burns and accidents(Deku et al., 2024). Of these, post surgical wounds are of the major burdens, causing surgical site infections(Zewdu et al., 2025). Data about bactaerial wound contamination is derived almost exclusively from studies in high-income countries yet the burden from is disproportionately higher in low- and middle income countries (Nyesiga, Nakibuuka, Kajumbula, Ssenyonga, & Byakika-Kibwika, 2022).

Globally, wound bacterial contamination is reported to be the third most common nosocomial infection preceded only by urinary tract infections and pneumonias (Marie Josée Mukagendaneza et al., 2019). The outcome of bacteria wound contamination is partly attributed to increase in antimicrobial resistant bacteria pathogens, which make the choice of empirical therapy more difficult(Deku et al., 2024; Shakir A, 2021). In low income countries, wound contaminations are costly and impose a heavy and potentially preventable burden on both patients and healthcare providers(Khan et al., 2023; Mehtar et al., 2020). The rates in East African countries have increased recently with Tanzania, Uganda, Ethiopia’s ranging from 21.4% to 59.29%(Ali, Neway, & Desta, 2025; Dawer et al., 2024; Weinreich et al., 2023).

In Uganda, data about wound contamination is still scarce and the true incidence and cost per patient are unknown. However, a study by Laura Jung et. Al., in 2023 at Mulago National Referral Hospital reported wound bacterial contamination to be as high as 98% (Jung et al., 2023). Moreover, a study by Khalim et al.,2021, reavealed that bacteria infecting wounds in Mbarara hospital were resistant to commonly used antibiotics(Khalim, Mwesigye, Tungotyo, & Twinomujuni, 2021), a scenario observed in other East African nations(Tshimbila Kabangu et al., 2026). Approximately ten additional days on the length of hospital stay have been attributed to wound bacterial contamination (Olowo-Okere, Ibrahim, Sani, & Olayinka, 2018). Records at Hoima Regional Referral Hospital (HRRH) showed that between July and August 2022, seven hundred and five (705) surgeries were performed and 168 had indications for surgical site infections with majority having treatment failure. In the microbiology register; 5 out of 10 wound swabs cultured showed positive cultures for organisms with multi drug resistance. This led to prolonged hospital stay and increased treatment costs for the patients as result of this ineffective treatment.

However, no study had been conducted to determine the prevalence of post operative wound bacterial contamination, bacterial isolates and their antimicrobial susceptibility in this low resource setting. This study therefore determined the prevalence of the common pathogenic bacteria associated with post operative wounds and determine the antibiotic susceptibility of the commonly used antibiotics. This will to improve patient management and reduce hospital acquired infections, reduce duration of stay in hospital and thus reduce unnecessary expenditure for treatment.

## 2.0 Materials and Methods

### 2.1 Study design

A cross-sectional study involving post-operative patients admitted in postnatal and surgical wards. The study was conducted for a period of 3 months, from the month of October 2022 to December 2022.

### 2.2 Study setting and participants

The study was conducted at Hoima Regional Referral Hospital (HRRH); located in Hoima district, western Uganda, bordering with the Democratic Republic of Congo (Magambo, Bajunirwe, & Bagenda, 2020). The Hospital serves five general government hospitals and a population of approximately 3,000,000 people; it lies approximately 200 kilometres north west of Kampala. The study partcipants included patients in the postnatal ward with ceasarean section wounds (188) and surgical ward with acute and chronic wounds (102) admitted to the hospital wards immidiately after surgical operation. The acute wounds included 60 accident patients, 15 burn patients, 3 appendectomies, 13 hania repairs and 5 patients from orthopedic procedures. Six were chronic wounds resulting from diabetic amputation procedures. These patients had stayed in the ward for 48hrs prior sampling and had already iniated onto prophylactic intravenous (IV) antibiotics immediately after surgery.

The study also took samples from the theatre surfaces including the air, door handles, equipment and floors.

### 2.3 Sample size and sampling procedure

A total of 290 samples from post operative wounds, 46 from theatre air and 28 from theatre surfaces were collected. Swabing followed a procedure described by Deborah Chester Bessell for obtaining a wound and theatre surface specimen for culture(Bessell, 2025). Briefly, a sterile swab was rotated from margin to margin in a 10 point zizgag fashion while exarting enough pressure to express enough fluid from the wound tissue. The swab was placed in the transport media and transported to the laboratory for analysis. Swabs were collected from patients 48hrs post admission and two swab samples were collected from each participant. Two hundred and ninety wound swabs were obtained from post-operative patients in Surgical, postnatal, orthopedic, and intensive care unit wards.

To determine the index of microbial air contamination in the theatre, 46 Hospital air samples (clean zone) were obtained from different sections of the threatre including; operating theatre rooms, sterile setting areas A and B, theatre reception, post anesthetic unit, theatre changing rooms (male and female rooms), sluice room and surgeon’s office following a procedure described by Mohammed O. A. Mohammed(Mohammed, 2023). The passive sampling involved placing Blood, Chocolate and MacConkey agar plates in the sampled rooms approximately 1m away from the operating table, windows and doors to avoid contamination. The plates were placed in duplicate and left open overnight.

### 2.4 Laboratory procedures

Swabs collected from wounds and surfaces were innoculated on duplicate plates of Blood Agar, Chocolate Agar and MacConkey Agar. Half of the plates were incubated aerobically and the other half anaerobically (sandwich method) for 18-48hrs at 37°C. For air samples, the plates were retrievd from the rooms and were examined for presence of colonies.

Bacterial colonies on the agar plates were gram stained and subjected to biochemical tests for identification and classification. Unclassified coliforms were lactose fermenting on MacConkey agar, yielding pink-coloured colonies, and were uniform gram negative rods. Antimicrobial susceptibility to commonly prescribed antibiotics at Hoima Regional Referral Hospital was determined following the Kirby – Bauer disc diffusion as previously described by Schiller *et al*.,(Schiller, Young, Schulze, Tripepi, & Pohlschroder, 2022). The antibiotics tested were; tetracycline 30µg, Trimethoprim sulfamethozale 1.25/23.75µg, ampicillin 10µg, gentamicin 10µg, impenem 10µg, ceftriaxone 30µg, ciprofloacin 5µg, piperacillin 100µg, cefdazidime 30µg, erythomcin 15µg, vancomycin 30µg, cefoxitin 30µg, Penicilin G 10 unints, and clindamycin 2µg. Resistance or susceptibility was interpreted following the Clinical Laboratory Standards (CLSI) version 2022. *E. coli* NCCP 13894, *P. aeruginosa* NCCP 12297 and *S. aureus* NCCP 11486 were used as quality control strains.

### 2.5 Data analysis

Data was entered in Microsoft Excel, sorted, cleaned and exported to SPSS version 23 (IBM Inc. USA). Data was described using frequencies, means and SDs. Chi square (χ2) test and Mann-Whitney U Test were used to determine differences in microbial growth patterns and antibiotic resistance patterns. A difference of *P < 0.05* was considered significant.

### 2.6 Ethical considerations

The study was approved by Institutional Ethical Review Committee of Mbarara University of Science and Technology. Written informed consent was obtained from participants ≥18 years. For participants < 18years, assent was got from their parents or caregivers. Eligible participants were free to decline participation and such action did not affect their care.

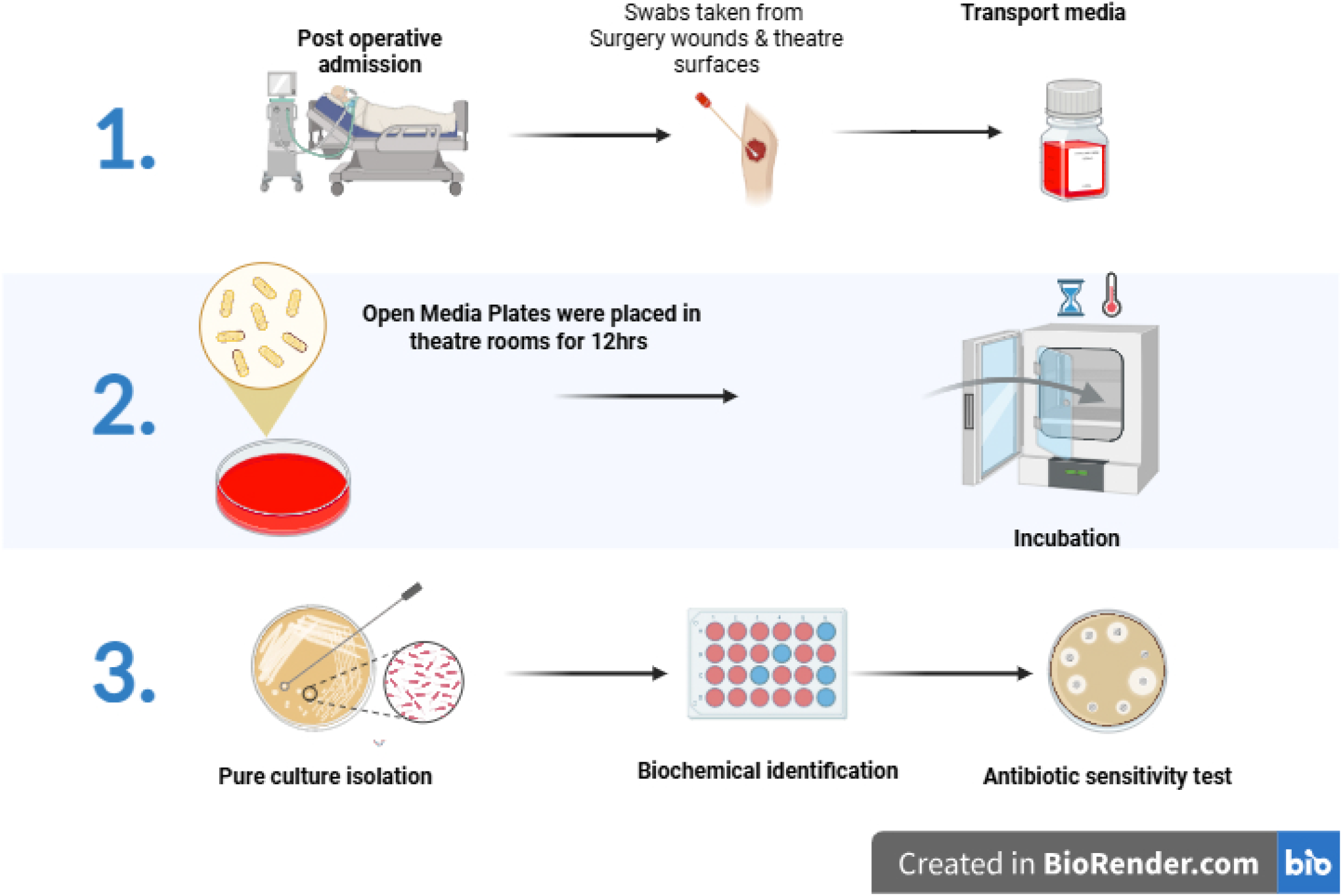
Graphical Abstract representing methodological approach undertaken. **1**, Post operative patients admitted to the hospital and after 24hr post admission, swabs are were taken. **2**, Sterile Media plates were placed in different theatre areas to collect determine air contamination. **3**, Pure cultures from wounds, theatre surfaces and air were isolated, gram stained and biomedically identified. The pure cultures were then tested for antibiotic sensitivity.

## 3.0 Results

### 3.1. Women of child bearing age were most admitted for management of Surgical site bacterial contamination

A total of 290 patients were followed up after surgery and wound swabs obtained from then. The demographic characteristics are summarised in Table 1. Majority of the patients, 188(64.8%) had been admitted to the postnatal ward and 102(35.2%) to the surgical ward. Most of the study participants were females 211(72.8%) while male participants accounted for 27.2%. Agewise, 14(4.8%) of the participants were less than 18 years, 116(40.0%) were 18 to 27 years, 93(32.1%) were 28 to 37 years, 43(14.8%) were 38 to 47 years and 24(8.3%) were 48 years and above. All the patients had been iniatiated on antibiotic therapy, either on a combination of Ceftriaxone and Metronidazole (84.9%), Ceftriaxone alone (7.9%), or piperacillin salbactam (PISA) (7.2%). Majority of the patients, 150 (51.6%) had spent 3 to 4 days on antibiotics, while 38.9% had spent more than 4 days and only 9.3% of the participants had spent 1 to 2 days on antibiotic treatment.

**Error! Reference source not found.**.

| Variable | Category | Frequency (n = 290) | Percentage (%) |
| --- | --- | --- | --- |
| Ward | Postnatal | 188 | 64.8% |
|  | Surgical | 102 | 35.2% |
| Gender | Male | 79 | 27.2% |
|  | Female | 211 | 72.8% |
| Age | Less than 18 | 14 | 4.8% |
|  | 18 to 27 | 116 | 40.0% |
|  | 28 to 37 | 93 | 32.1% |
|  | 38 to 47 | 43 | 14.8% |
|  | 48 years and above | 24 | 8.3% |
| Admitted in past 24hrs | Yes | 290 | 100.0% |
|  | No | 0 | 0.0% |
| Patient been on antibiotics in past 24hrs | Yes | 290 | 100.0% |
|  | No | 0 | 0.0% |
| Antibiotics | Ceftriaxone alone | 23 | 7.9% |
|  | Ceftriaxone and Metronidazole | 246 | 84.9% |
|  | Pisa | 21 | 7.2% |
| Duration on antibiotics | 1 to 2 days | 27 | 9.3% |
|  | 3 to 4 days | 150 | 51.6% |
|  | More than 4 days | 113 | 38.9% |

### 3.2 Surgical site/wound contamination was significantly associated with age, gender and duration of antimicrobial therapy

There was microbial growth on 89(30.7%) of samples isolated from the wounds of patients admitted for surgical site infection management (Figure 1A). Although majority of patients with SSIs were from the postnatal ward (32.9%), there was no significant difference with SSIs in patients from surgical ward (26.5%) (χ^2^= 1.1317, df=1 *P=0.2512) (Figure 1A)*. We further observed that SSIs was significantly higher in females (85.4%) as compared to males (14.6%)(χ^2^= 10.79, df=1, *P=0.0010) (Figure 1B)*. Furthermore, there was a significant difference in SSIs based on age groups (Figure 1C). Young adults aged 18-37 years had higher prevalences of SSIs ranging from 31.9% to 38.7% as compared to the individuals aged less than 18 years and those aged aboved 38 years (χ^2^= 18.38, df=4, *P=0.0010)*. Although, microbial growth was higher in individuals receiving Ceftriaxone alone (34.8%), it was not significantly different from individuals receiving PISA or a combined therapy of Ceftriaxone and Metronidazole(χ^2^= 4.839, df=2, *P=0.0890*) (Figure 1D). Lastly, we observed that duration on antibiotic treatment influenced microbial growth in individuals suffering from SSIs (Figure 1E). Microbial growth increased with an increase in duration of antibiotic therapy, with least number of SSI patients who had received antibiotic drugs for 1-2 days having less microbial growth (11.1%) as compared to individuals who had received for more than 4 days (41.6%) (χ^2^=12.73, df=2, *P=0.007*). These results suggest that gender, age and duration of antimicrobial therapy can influence microbial growth in patients suffering from SSIs.

**Figure 1.**
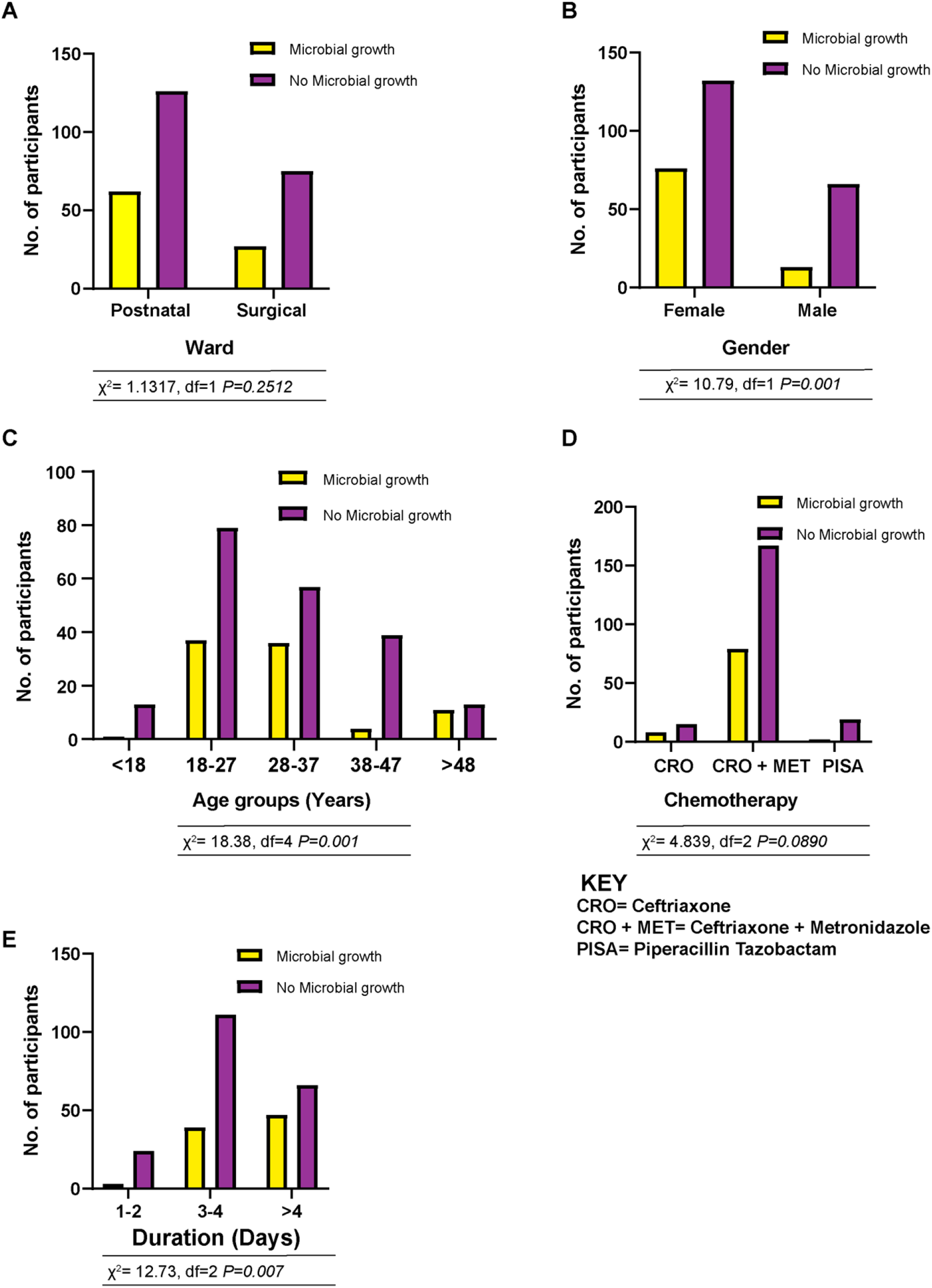
Prevalence of surgical site/wound contamination Among post – operative patients in Hoima Regional Referral Hospital Referal Hospital. **A**, Prevalence of surgical site/wound contamination based on ward of admission; **B**, Gender; **C**, Age; **D**, Antimicrobial therapy; **E**, Duration of therapy. Comparasion among different groups was estimated using Chi square. The χ^2^ value, df and *P*-values are given under each respective figure.

### 3.3 Gram negative bacteria are the major causes of Surgical Site Contamination among post-surgical patients at Hoima Regional Referral Hospital

A total of 115 isolates were grown from wounds among post surgical patients. These organisms were grouped as gram positive and gram negative organisms. The gram negative bacteria isolated (87) were higher than the gram positive bacteria(28), although this was not statistically significant (Figure 2A). The isolated gram gram negative bacteria belonged to seven (7) genera while the gram positive belonged to 3 genera. Of the total isolates cultured, *E.coli* was the most prevalent (32.2%), followed by *Staphylococcus aureus* (13.9%) as shown in Figure 2B. *Acinetobacter* spp and *Klebsiella pneumoniae* had a prevalence of 11.3%, while *Enterobacter* spp (7.5%), *Pseudomonas* spp (5.2%), *Proteus spp* (4.3%), *Citrobacter* spp (3.5%). Our findings suggest that gram negative bacteria, mostly *E. coli* are the most causative agent of SSIs among post operative patients admitted at Hoima Regional Referral Hospital Regional Referal Hospital.

**Figure 2.**
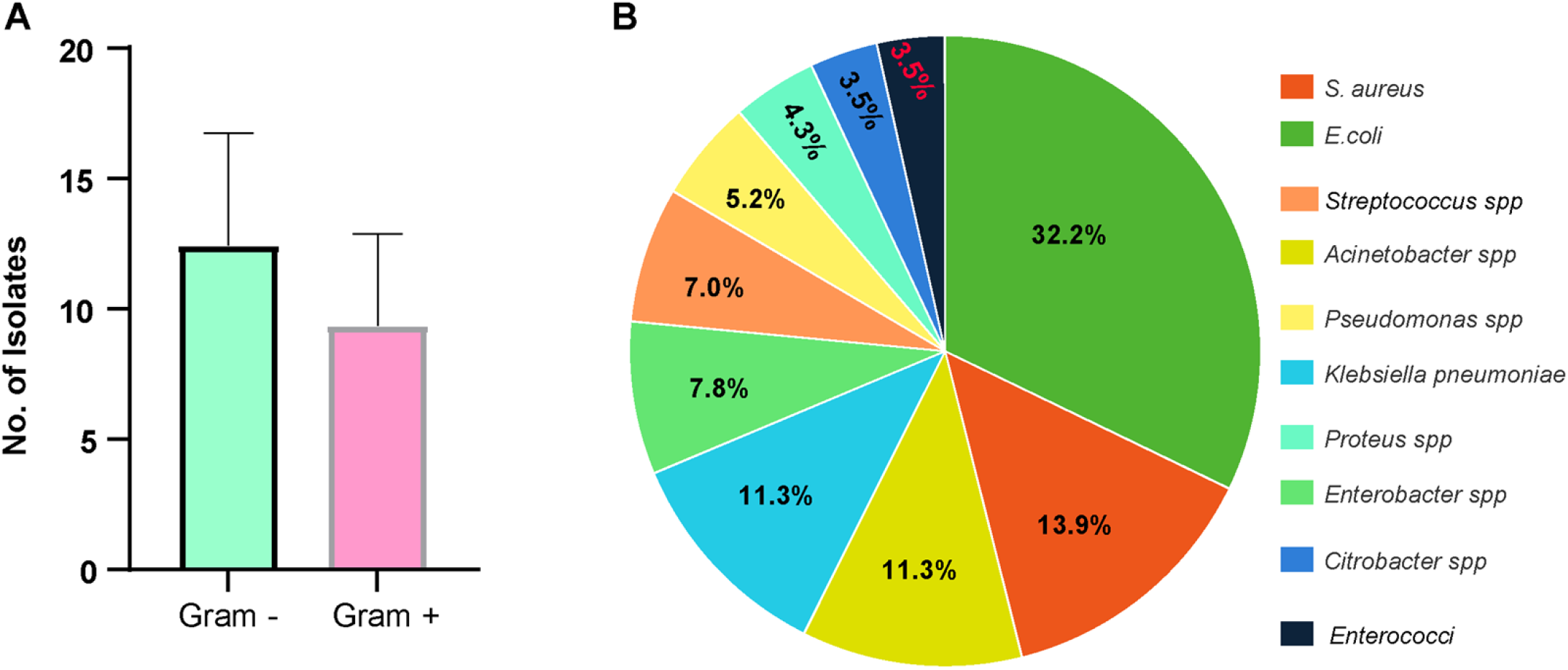
Bacterial spp involved in SSIs among post surgical patients. **A**, Isolated bacteria grouped based on their gram staining characteristics. **B**, Prevalence of various bacterial spp isolated from septic wounds among post surgical patients admitted at Hoima Regional Referral Hospital.

### 3.4 Air space contamination did not posse a significant risk as a source of microorganisms involved in Surgical Site/wound contamination

Overall, 49 isolates were obtained from air and surgical surfaces in Hoima Regional Referral Hospital. Air spaces had significantly higher isolates as compared to theatre and ward surfaces (Figure 3A). Fourty three (86%) isolates were isolated from air spaces of different rooms while 6 isolates were cultured from different surfaces of theatre and wards. Among the surfaces, the sanction machine had higher percentage of isolates (42.8%), followed by the floor (28.6%) while the sinks and trolley surfaces each had 14.3% of the total surface isolates (Figure 3B). The major organisms that were isolated from surfaces and air spaces included Coagulase Negative *Staphylococcus aureus* (CNS) and *Corynebacterium* spp (COR). Air spaces were significantly contaminated with CNS as compared to COR while the surfaces were significantly contaminated with COR compared to CNS (Figure 3C) (χ^2^=9.090, df=1, *P=0.0026*). Air space being significantly contaminated could point to poor space sterilization procedures or faulty structural engineering, exposing the theatre spaces to pathogens.

**Figure 3.**
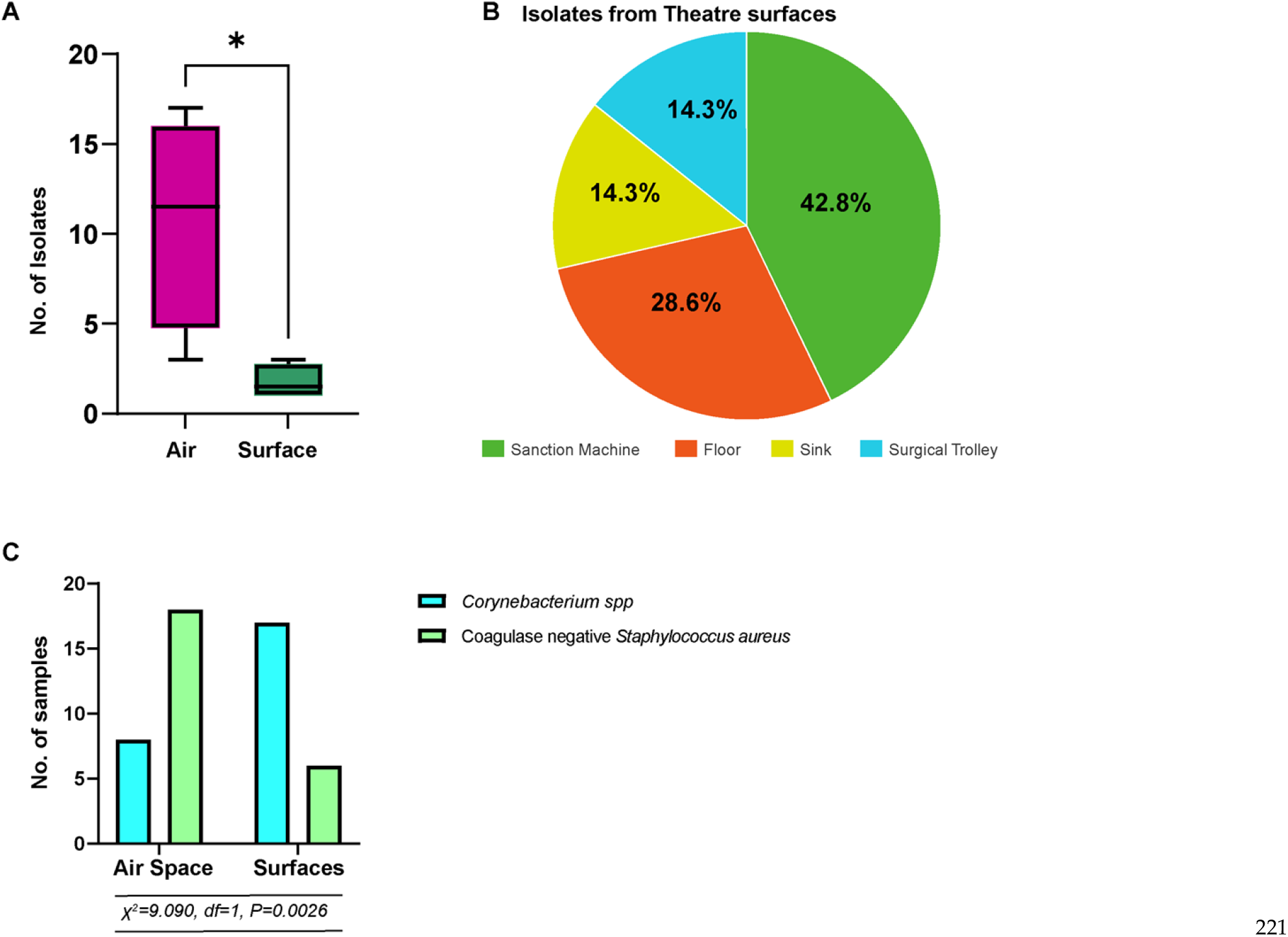
Contamination of surgical air space and surfaces at Hoima Regional Referral Hospital. **A**, Number of isolates cultured from air space and surfaces. Comparasion among different groups for number of isolates was estimated using unpaired t-test. **B**, Number of isolates obtained from different surfaces. **C**, Genera of bacteria isolated from air spaces and surfaces. Comparasion among different groups for genera was estimated using Chi square. The χ^2^ value, df and *P*-values are given under the figure. * < 0.05

### 3.5 Pathogens Isolated from Surgical wounds and theatre surfaces showed multidrug resistance

We performed the KB antibiotic susceptability testing on the most prevalent bacteria isolated from wounds, theatre air spaces and surfaces. Duplicate samples were analysed for resistance using the 2-Way ANOVA. *E.coli, Klebshella, Acinetobacter and Enterobacter* were tested for antimicrobial reistance. The results revealed a significant difference in resistance among these isolates *(P<0.001)* (Figure 4). Resistance to Septrine and Ceftriaxone was higher (80-100%) among all isolates, while least resistance (0-20%) was towards Imipenem, with only *E.coli* (2.7%) and *Acinetobacter* (15.4%) showing resistance to Imipenem. Also, resistance among the different microorganisms was significantly different *(P<0.001)*, as high reistance was seen among *E.coli* and *Acinetobacter* isolates as compared to *Enterobacter* and *Klebshella* (Figure 4). The reistance to all antibiotics was observed among the former while the later were 100% sensitive to imipenem. On the other hand, 57.1% and 25% of *Staphylococcus aureus* were resistant to erythomycin and penicillin respectively. However, all *S. aureus* strains were 100% sensitive to Ciprofloxacin, Trimethoprine and Tetracycline. *Corynabacterium* was sensitive to all antibiotics tested. Our results suggest that most organisms isolated from septic wounds were multidrug resistant.

## 4. Discussion

Wound contamination remain a public health threat especially in low resource settings leading to prolonged hospital stays and associated burden of diseases. In this current study, we assessed the prevalence of post operative wound bacterial contamination, associated bacteria isolates and determined their antimicrobial susceptibility patterns towards commonly prescribed antibiotics at Hoima Regional Referral Hospital. Our study found that, the proportion of patients with bacterial wound contamination was 30.7%, lower than national prevalence reported at 47.1%(Lubega, Joel, & Justina Lucy, 2017). However, this was comparable to other studies in East African countries, where wound contamination range from 21.4% to 31.8%. A 10.9% incidence was reported in Bugando Medical Centre of Tanzania (Mpogoro et al., 2014), while Atul Singh reported a 92.31% of microbial growth at a tertiary hospital in India among post operative patients(Singh, Sharma, & Kumar, 2025). The high prevalences of bacterial wound contamination in low income countries could be due to lack of resources, poor committement of government to better health services and low economic status of majority of the population (R. E. Mengesha, B. G.-S. Kasa, M. Saravanan, D. F. Berhe, & A. G. Wasihun, 2014). These alarming high prevalences are contrally to what is observed in developed countries, with low wound contamination, ranging from 2-5% (Sandy-Hodgetts et al., 2022). This contrast is related to the variations in infrastructural and sterilisation facilitities, standard infection control practices, and human resource capacity, which are more standardised in developed nations.

**Figure 4.**
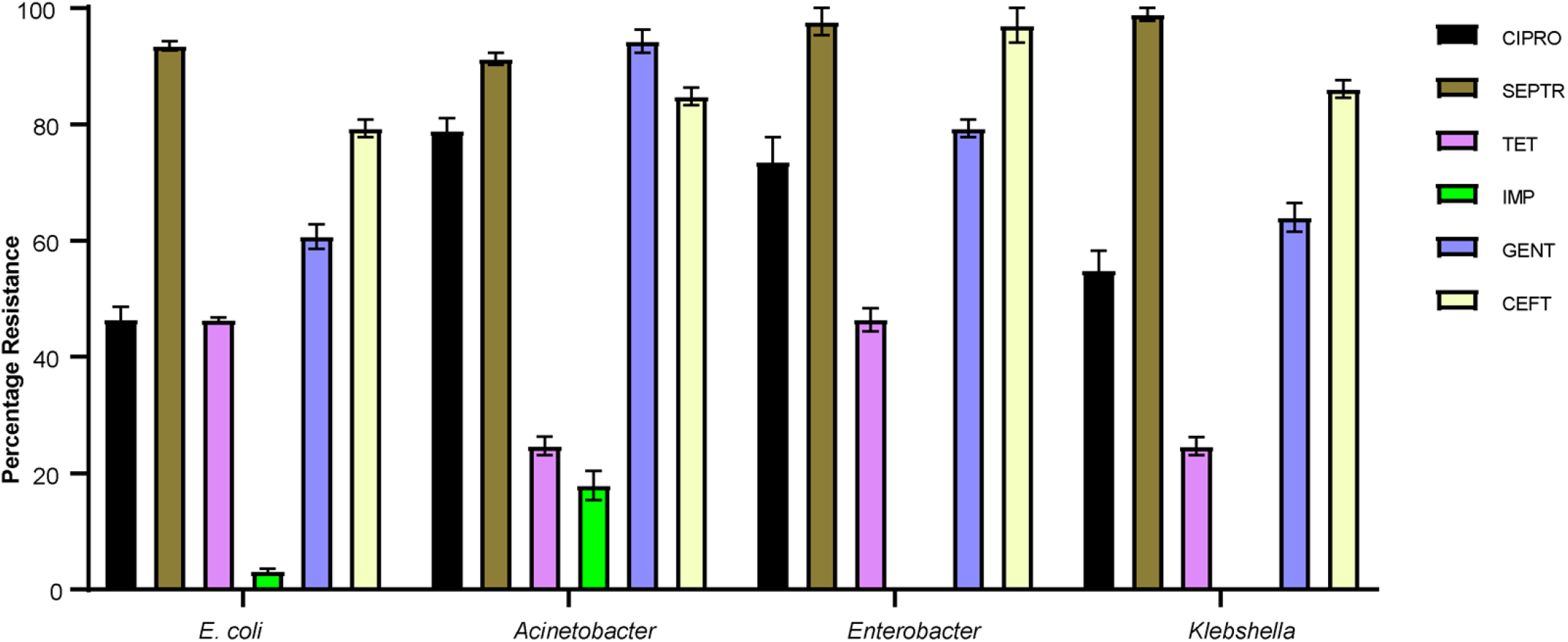
Prevalence of resistance to commonly prescribed antibiotics among bacteria isolated from septic wounds among post operative patients and theatre surfaces at Hoima Regional Referral Hospital. Results were analysed in duplicate using 2-way ANOVA and data is graphically represented using SEM. (CIPRO= Ciproflaxicin, SEPTR= Septrin, TET= Tetracycline, IMP= Imipenem, GENT= Gentamycin, CEFT= Ceftrioxone)

Furthermore, females and young adults aged 18-37 years had a significantly higher prevalence of bacterial wound contamination of upto 85.4% and 38.7% respectively. This was related to the high proportion of postnatal cases within the sample population. Moreso, it is reported that obstetric and gynecological surgeries including cesarean deliveries are major risk factors of bacterial wound contamination in low-resource settings(WHO, 2020). Similar scenarios have been reported at Mbarara Regional Referral Hospital where the prevalence was 7.9% in young mothers aged 25 years (Mwandah et al., 2024). A retrospective study in Kenya by David Odada also reported a 2.5% of prevalence in mothers following cesearing deliveries(Odada, Shah, Mbithi, & Shah, 2024).

Our study further found out that, despite prophylatic antimicrobial therapy initiated post surgical operation in the patients, there was microbial growth. This was highest among individuals receiving Ceftriaxone alone, which is the most prescribed drug in Ugandan health facilities post operative procedures(Kutyabami et al., 2021). However, its also the most abused drug in the country (Edith, Mpirirwe, & Kiguba, 2024), risking development of antibiotic resistance. Increased microbial growth and prolonged antibiotic exposure suggests selective pressure favoring resistant organisms responsible for hospital acquired infections, which is reported in African healthcare facilities(Tadesse et al., 2017).

Gram negative bacteria were the most predominant clinical isolates, wih *E. coli* (34.9%) being the most common pathogen responsible for the highest cases of post operative wound contamination. Similar findings have been reported by other studies in East Africa, where enteric pathogens have been commonly associated with wound contamination (R. E. Mengesha, B. G. Kasa, M. Saravanan, D. F. Berhe, & A. G. Wasihun, 2014; M. J. Mukagendaneza et al., 2019). *E. coli* was the predominat cause of wound contamination due to the posibility of endogenous flora contamination, poor hand hygiene and sanitation; breaches in aseptic technique during the surgical procedure (Bucataru et al., 2023; Harrop et al., 2012). *Staphylococcus aureus* accounted for 15.1% of wound contamination cases and the least proportion of contamination was caused by *Pseudomonas* spp. (0.9%). These findings are contrary to those reported by George and colleagues, who reported *Staphylococcus aureus* as the most common organism (39.4%) contaminating wounds (George, Iramiot, Muhindo, Olupot-Olupot, & Nanteza, 2018). The difference in findings could be related to a difference in study settings.

Theatre surface sampling revealed that air spaces were the most contaminated reservoir of bacteria and it accounted for 86% of the total isolates. This is in line with previous studies that have shown that airborne contamination in operating rooms is the major contributor of bacterial wound contamination (Seth Caous et al., 2025; Stauning et al., 2020). Among the bacteria isolated from the surfaces, CNS was more prevalent (60.0%), suggesting contamination from the skin flora. These findings are comparable to those at Mbale Regional Referral Hospital in a study by (Reiye Esayas Mengesha et al., 2014), who found that 14.6% of the total isolates among post operative patients were CNS. Also, a study by Himani Kumari reported CNS to be the most predominant bacteria isolated from the floor of a university hospital in India(Kumari, Chakraborti, Singh, Chakrawarti, & Mukhopadhyay, 2020).

Antimicrobial susceptibility testing showed a high resistance to some of the commonly used antibiotics including Trimethoprim - sulfamethoxazole, Ceftriaxone, Gentamicin, and Ciprofloxacin among all gram negative isolates. Antimicrobial resistance was higher amongst *E.coli* and *Acinetobacter* isolates compared to Klebshella and Enterobacter. These findings were contrally to a smillar study done at Mbarara Hospital by Derrick Hope, who reported Klebshella to be the most resistant bacteria to antimicrobials (Hope et al., 2019). Also, our findings revealed that only *E.coli* and *Acinetobacter* showed some resistance to Imipenem, a finding smillar to a study by Refath Farzana who reported resitance to Imipenem in Acinetobacter among clinical isolates in Bangaladesh and a Jilin hospital respectively(Farzana, Swedberg, Giske, & Hasan, 2022; Roberts, Hassan, Li, & Li, 2016). Generally, Gram positive isolates were sensitive to most tested antimicrobials especially Ciprofloxacin, Trimethoprine and Tetracycline. However, we noted CNS resistance to erythomycin and penicillin. This aligns with Atul Singh’s results who found 33.33% of *S. aureus* isolated from post operative wounds to be resistant to commonly prescribed antibiotics (Singh et al., 2025).

## 5. Conclusions

Overall, our study demonstrated that bacterial wound contamination and antimicrobial reistance in HRRH is high. We recommend strengthening of infection control practices to prevent wound contamination. Right antibiotics for post operative patients should be prescribed following recommended procedures.

This study had some limitations, including the small number of isolates and the short duration of the study. In addition, performing genotypic resistance studies would have indicated the origin and type of resistance amongst these isolates. Despite these limitations, our study contributes to prevention of wound contamination at HRRH and in low resource settings.

## Data Availability

NA

## Ethics Statement

The study was approved by the Institution Review Board at MUST and presented to the Director and in-Charge of the Laboratory department at Hoima Regional Referral Hospital who granted permission to conduct the study. The study also followed the ISO 14644-1:2015 standard operating procedures. Informed consent was obtained from all subjects involved in the study.

## Acknowledgments

We thank the coordinator Medical Laboratory Sciences – MUST, Lecturers and non-teaching staff at MUST – Mulago Campus and all BMLS year three students for all support rendered throughout the research study. We sincerely acknowledge the efforts of Mr. Buga Rudolf, our supervisor, who guided us throughout the time of conducting the research study. We are indebted to the administration and staff at Hoima Regional Referral Hospital, who granted permission, offered assistance and guidance to us for the success of the research study. We are indebted to the research participants for their consent to participate in this research study.

## Conflicts of Interest

The authors declare no conflict of interest.

## Author Contributions

**Conceptualization**; Abarteneko Chelangat and Jimmy Omeke, **Methodology;** Moses Tabule Musanad Asimai, Abarteneko Chelangat, **Validation;** John Roberts Padde, Winnie Akiteng, **Formal analysis;** John Roberts Padde, Abarteneko Chelangat, Resources, Rudolf Buga, Bwaga Ibrahim, **Data curation;** John Roberts Padde, Abarteneko Chelangat, **Writing original draft preparation;** Abarteneko Chelangat, Jimmy Omeke, John Roberts Padde, Musanad Asimai, **Review and editing;** John Roberts Padde, Winnie Akiteng, Abarteneko Chelangat.

## Funding

This research received no external funding.

## Notes

### Competing Interest Statement

The authors have declared no competing interest.

### Clinical Trial

NA

